# A Specialized Reference Panel with Structural Variants Integration for Improving Genotype Imputation in Alzheimer’s Disease and Related Dementias (ADRD)

**DOI:** 10.1101/2024.07.22.24310827

**Authors:** Po-Liang Cheng, Hui Wang, Beth A Dombroski, John J Farrell, Iris Horng, Tingting Chung, Giuseppe Tosto, Brian W Kunkle, William S Bush, Badri Vardarajan, Gerard D Schellenberg, Wan-Ping Lee

## Abstract

We developed an imputation panel for Alzheimer’s disease (AD) and related dementias (ADRD) using whole-genome sequencing (WGS) data from the Alzheimer’s Disease Sequencing Project (ADSP). Recognizing the significant associations between structural variants (SVs) and AD, and their underrepresentation in existing public reference panels, our panel uniquely integrates single nucleotide variants (SNVs), short insertions and deletions (indels), and SVs. This panel enhances the imputation of disease susceptibility, including rare AD-associated SNVs, indels, and SVs, onto genotype array data, offering a cost-effective alternative to whole-genome sequencing while significantly augmenting statistical power. Notably, we discovered 10 rare indels nominal significant related to AD that are absent in the TOPMed-r2 panel and identified three suggestive significant (p-value < 1E-05) AD-associated SVs in the genes *EXOC3L2* and *DMPK*, were identified. These findings provide new insights into AD genetics and underscore the critical role of imputation panels in advancing our understanding of complex diseases like ADRD.

## Introduction

Genome-wide association studies (GWAS) aim to identify genomic variants linked to disease risks or specific traits by analyzing the genomes of numerous individuals. GWAS seeks to identify variants that occur more frequently in individuals with a particular disease compared to those without it. GWAS primarily employs either whole-genome sequencing (WGS) or genotyping arrays to identify genomic variants. Despite the rapid advancements and increasing affordability of WGS technology, it still remains prohibitively expensive and computationally demanding for large-scale cohorts. Consequently, genotype arrays provide a pragmatic and valuable tool due to their cost-effectiveness and the availability of extensive disease data.

Genotype arrays assay variants relying on a pre-designed set of a small fraction of variants chosen by the linkage disequilibrium (LD) structure of the human genome. Variants not directly genotyped on arrays can be statistically inferred through a process called genotype imputation, which compares variants in haplotypes to an external reference panel containing known haplotypes of a large number of individuals, who have been genotyped using high-density genotype arrays or WGS. Usually, imputation algorithms first estimate haplotypes between each individual in a study cohort utilizing genotype arrays and a reference panel, and then use this information to infer missing alleles of the individual. The accuracy of imputation depends on several crucial factors, including haplotype size, the accuracy of genotypes in individuals, and the population diversity of the reference panel.

Currently, several public reference panels exist, such as the International HapMap Project^1^, the 1000 Genomes Project (1000GP)^2^, the UK10K Project^3^, the Haplotype Reference Consortium (HRC)^4^, and the Trans-Omics for Precision Medicine (TOPMed) program^5,6^. Among these, the TOPMed-r2 panel stands out with its reference panel including 97,256 WGS samples, making it the largest reference panel for genotype imputation to date^6^. The most recent version, TOPMed-r3, was released in December 2023. However, during the experiments conducted for this study, only TOPMed-r2 was available.

While public reference panels demonstrate high imputation accuracy in European populations, their effectiveness is limited when applied to other ethnicity groups. Population-specific reference panels, such as those tailored to Asian^7^ and African^8^ populations, show improved performance by capturing recently evolved population-specific variants. Similarly, public reference panels, composed of common populations, may potentially neglect rare variants in particular diseases. Therefore, we hypothesize that utilization of disease-specific imputation panels may improve imputation accuracy for disease studies.

Another rationale for the necessity of disease-specific imputation panels is that current public reference panels either lack or have a limited number of structural variants (SVs) that have been implicated in the association with human diseases^9,10^. For example, the association of the inverted H2 haplotype with reduced risk of a range of neurodegenerative diseases^11^, an 18⍰Kb copy number variation in *CR1* was found to associate with AD^12^ and an 8 kb deletion upstream of *CREB1* is also associated with AD^13^. Recent advancements underscore the importance of SV imputation. One study developed a multi-ancestry SV imputation panel using long-read sequencing data of 888 samples from 1000GP^14^, and another study on the *CYP2A6* gene emphasized genotyping and imputing known and novel SVs to understand genetic influences on traits like nicotine metabolism^15^. These findings illustrate that incorporating SVs into imputation panels enhances the resolution and accuracy of genetic association studies, providing deeper insights into the genetic underpinnings of complex diseases such as AD. By capturing a broader spectrum of genetic variation, including SVs, disease-specific imputation panels offer a more comprehensive tool for genomic research, facilitating better disease risk prediction and understanding of disease mechanisms.

The Alzheimer’s Disease Sequencing Project (ADSP) is a collaborative research effort, sequencing diverse individuals across populations. The ADSP Release 3 (R3) 17K contains 16,905 samples with WGS data. Leveraging data from ADSP, we built ADSP-Short-Var (single nucleotide variants [SNVs] and short insertion/deletions [indels]) and ADSP-All-Var (SNVs, indels, and SVs) reference imputation panels, tailored to capture AD-enriched variants, particularly for SVs. We demonstrated the strengths of these specialized panels by applying them to genotype data of 38,271 subjects of multiple ethnicities from the Alzheimer’s Disease Genetics Consortium (ADGC).

## Results

### Overview of ADSP-Short-Var, ADSP-All-Var, and TOPMed-r2 Panels

The ADSP-Short-Var panel contained 54 million variants (51,459,037 [93.94%] SNVs and 3,322,380 [6.06%] indels in chromosomes 1-22) derived from 16,564 sequenced genomes, representing diverse ethnic backgrounds including 62.76% non-Hispanic white, 18.68% Hispanic, 18.11% African American, 0.3% Asian, and 0.14% other ethnicities. In comparison, the TOPMed-r2 panel included 295 million variants (274,388,520 [92.85%] SNVs and 20,899,436 [7.15%] indels in chromosomes 1-22) derived from 97,256 sequenced genomes, 48.49% European, 24.95% African, 17.57% admixed American, 1.22% East Asian, 0.66% South Asian, and 7.11% unassigned-ethnic individuals (**Table S1**). We categorized variants into three categories for comparison: consensus imputed variants shared between the imputations generated against the ADSP-Short-Var and TOPMed-r2 panels, ADSP-Short-Var-specific imputed variants, and TOPMed-r2-specific imputed variants. Among these variants, 50 million consensus variants were shared between the two panels (94.86% SNVs and 5.14% indels; **Figures 1A-B**). Panel-specific variants included 4 million variants (82.41% SNVs and 17.59% indels) unique to the ADSP-Short-Var panel and 241 million variants (92.42% SNVs and 7.57% indels) unique to the TOPMed-r2 panel.

**Figure 1.**
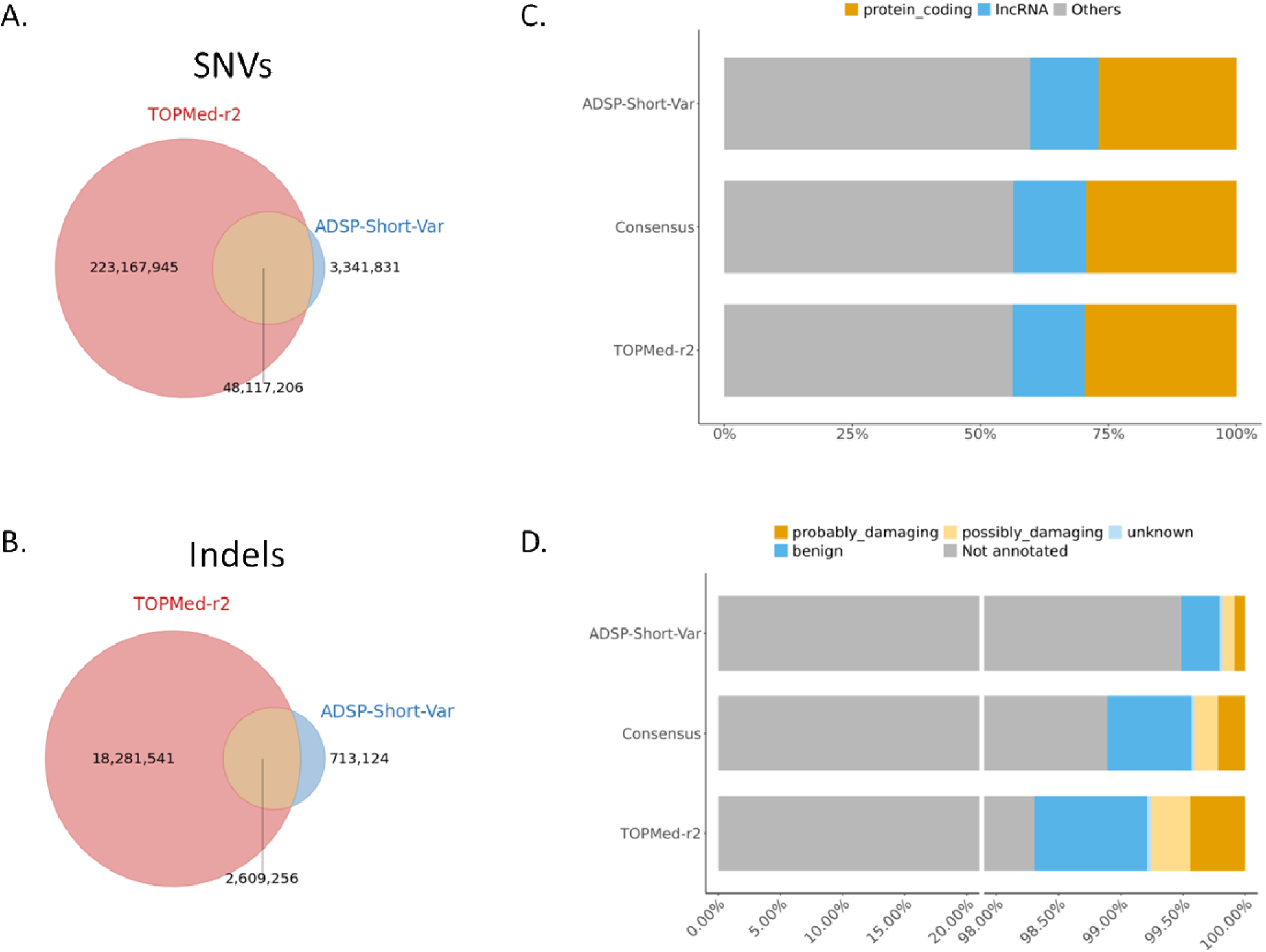
Comparison of variants between TOPMed-r2 and ADSP-Short-Var panel. (**A**) Venn diagrams showing the number of Single Nucleotide Variants (SNVs). (**B**) Venn diagrams showing the number of insertions/deletions (Indels). (**C**) Distribution of annotated biotypes. (**D**) PolyPhen predictions for TOPMed-r2 specific, ADSP-Short-Var specific, and consensus variants.

Using Variant Effect Predictor^16^ (VEP v105.0) for annotation, 29.32% of consensus variants were situated in protein-coding regions. For ADSP-Short-Var-specific and TOPMed-r2-specific variants, the percentages were 26.98% and 29.64%, respectively (**Figure 1C**). The second most common biotype observed was Long Non-Coding RNA (LncRNA), accounting for 14.26%, 13.38%, and 14.04% of consensus, ADSP-Short-Var-specific, and TOPMed-r2-specific variants, respectively (**Figure 1C**). Regarding variants with PolyPhen, the prediction labeled by possibly damaging and probably damaging, we observed 0.41%, 0.18%, and 0.76% in consensus, ADSP-Short-Var-specific, and TOPMed-r2-specific variants, respectively (**Figure 1D**).

The ADSP-All-Var panel was constructed by integrating SNVs and indels in the ADSP-Short-Var panel along with the SVs (231,385 deletions, 119,648 insertions, 45,839 duplications, and 3,362 inversions) identified on the same sample set in our previous study^17^. This integration enriched a diverse genomic landscape in the ADSP-All-Var panel, with proportions of 93.25% SNVs, 6.02% indels, 0.42% deletions, 0.22% insertions, 0.08% duplications, and 0.01% inversions, respectively. The incorporation of SVs into the ADSP-All-Var panel enables a more comprehensive discovery of variants associated with AD on large sample sets with genotype data.

### Discovery of Novel Suggestive Significant and Disease Susceptibility SVs Through Imputation

In an effort to enhance the statistical power of SV analysis, we performed imputation on the ADGC genotype dataset (Ncase=16,779 and Ncontrol=21,492) against the ADSP-All-Var panel. By increasing the sample size, we aimed to uncover novel significant SVs. Our subsequent single variant association test on the imputed SVs revealed three suggestively significant (p-value < 1E-05) SVs (**Figure S1**).

The most notable discovery was an Alu insertion in the intron of *EXOC3L2*, exhibiting a p-value of 1.78E-07, with allele frequencies (AF) of 0.01632 in AD cases and 0.01061 in controls. The further experimental validation by PCR also confirmed this insertion (**Figure S2**). This insertion is also present in the gnomAD database with an AF of 0.01275, similar to the AF of controls in our dataset. Another significant SV identified was a deletion at chr19:45775716 (p-value = 9.94E-06) located in intron 8 of *DMPK*. However, this region is complex, containing multiple Alu elements, such as AluSx1, AluJo, AluSz, AluSx3, AluY, and AluJb, which may affect the quality of the deletion call. It is crucial to note that the significance of the insertion at chr19:45216933 and the deletion at chr19:45775716 diminished under the condition of *APOE e4*, suggesting the confounding impact of the SVs and APOE *e4*. **Table 1** provides detailed information on these findings.

**Table 1.**
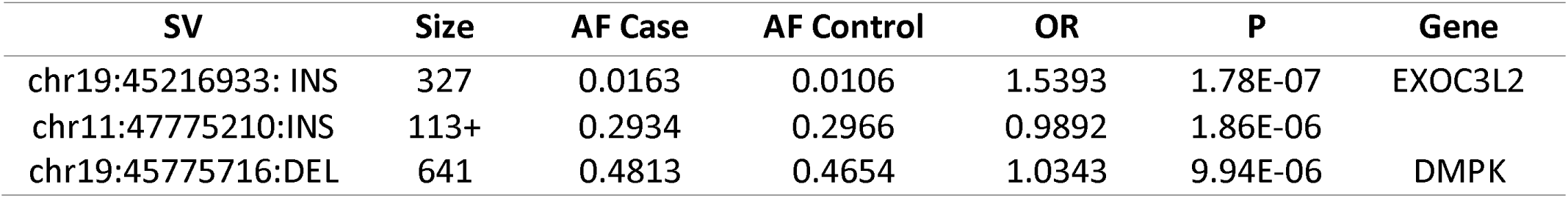
Three suggestive significant structure variants imputed by the ADSP-All-Var panel.

In the previous study on ADSP R3 17K WGS^17^, 107 SVs (72 deletions, 20 duplications, and 15 insertions) were reported. We found that 97.20% (104 out of 107) SVs were successfully imputed by ADSP-All-Var panel. With this larger sample set, 65.38% (68 out of 104) SVs exhibited an increase in allele count, and 39.71% (27 out of 68) showed enhanced statistical significance (**Table S2**). All the well-imputed SVs had highly similar AFs (r = 0.9931) to SVs discovered from ADSP (**Figure S3**).

Among these SVs, some were specifically located in important AD genes and were in the same LD block with known SNVs, which facilitated quality of the SV imputation of SVs. For instance, a 5,505bp deletion at chr2:105731359 in the upstream of *NCK2* (R^2^ = 0.7), which is a gene highly expressed in amyloid-responsive microglial cells, was in the same LD block with the known SNV rs143080277, associated with late onset AD^18^. Similarly, a 238bp deletion at chr17:46009357 in *MAPT* (R^2^ = 0.98), which encodes tau protein implicated in AD pathology, was in the same LD block with the SNV rs8070723, which is associated with reduced risk of LOAD^11^. This 238-bp deletion, located between exons 9 and 10 on the H2 background, is commonly used to differentiate between H1 or H2 haplotypes.

### Discovery of Disease Susceptibility SNVs and indels Through Imputation

In the analysis of WGS data from the ADSP R3 NHW cohorts (Ncontrol = 2,601 and Ncase = 4,053), 69 exonic rare indels (MAF < 1%) suggestively associated with AD were identified. These indels met the criteria with CADD > 20, p-value < 0.05 (Fisher’s test), and odds ratio (OR) exceeding 1.5 or under 0.5. Out of these 69 indels, 55 were confirmed through experimental validation. Given their presence in our ADSP-Short-Var panel, we imputed these indels on genotype data from the ADGC NHW cohorts (Ncontrol = 15,216 and Ncase = 13,182) to enhance statistical power by increasing sample size. As a result, the nominal significance of a deletion, chr20-663684-CCGGCGGGGGT-C in the exon 2 of *SCRT2*, increased with p-value from 0.0096 to 0.0034. *SCRT2* is a neuron-specific gene involved in neuronal survival, neuronal migration, and neurogenesis during brain development^19–21^. A study indicated that the SCRT2 expression was altered after surgery in aged mice with impaired cognition^22^. Of note, 10 out of 55 indels were absent in TOPMed-r2 imputation (**Table S3**), which are in genes, *C12orf81*, *TOMM20L*, *FAM174B*, *NTN3*, *RGL3*, *PNKP*, *PPDPF*, and *PCDHB13*.

To evaluate the accuracy of imputed genotypes for those disease susceptibility indels, PCR validation was conducted on six indels, which are located in *DNAH14*, *ANO7*, *ZNF655*, *PTGER1*, *SCRT2*, and *PPDPF*, on 17 available DNA samples from the ADGC cohorts (**Table S4**). The results revealed that 86.67% (13/15) indels were accurately genotyped by the ADSP-Short-Var panel, while only 66.67% (10/15) were accurately genotyped by the TOPMed-r2 panel.

We also investigated the 263 SNVs and 10 indels in *ABCA7*, previously discovered through the association test of aggregate of rare coding variants^23^ on the WGS data of ADSP R3 NHW cohorts. Among these variants, when imputed them onto ADGC NHW genotype data, 23.81% (65 out of 273) were well imputed by the ADSP-Short-Var panel, while 76.19% (208 out of 273) could not be imputed due to their rarity (AC < 5). For those well-imputed SNVs and indels, 75.38% (49 out of 65) showed an increase in allele counts as sample size expanded, and 67.35% (33 out of 49) increased statistical significance (**Table S5**). Importantly, all well imputed SNVs and indels had similar AFs (r= 0.9375) compared to AFs discovered from the WGS data of ADSP R3 NHW cohorts (**Figure S4**).

### Assessment of Imputation Accuracy of SNVs and Indels Compared to WGS

To evaluate imputation accuracy, we compared genotypes of SNVs and indels derived from imputations with those obtained from WGS. This analysis was conducted on a dataset with 2,363 Non-Hispanic White (NHW), 2,191 Hispanic, 799 African American (AA), and 43 Asian individuals with both genotype and WGS data available. Notice that these samples were independent from the ADSP-Short-Var and ADSP-All-Var panels.

Using aggRsquare^24^, we reported an aggregated R^2^, the squared Pearson correlation between genotypes obtained from imputations and WGS, across all SNVs and indels, stratified by minor allele frequency (MAF) bins: <0.0005, 0.0005-0.001, 0.001-0.005, 0.005-0.01, 0.01-0.05, and >0.05. The ADSP-Short-Var panel demonstrated improved performance over the TOPMed-r2 panel for variants with MAF < 0.0005 in Hispanic cohorts and performed comparably well for variants with MAF < 0.0005 in NHW cohorts (**Figure 2, Table S6**). Due to limited sample sizes, we were unable to examine the variants with MAF < 0.005 for AA and Asian cohorts. Conversely, the TOPMed-r2 panel outperformed for variants with MAF > 0.005 (**Figure 2**).

**Figure 2.**
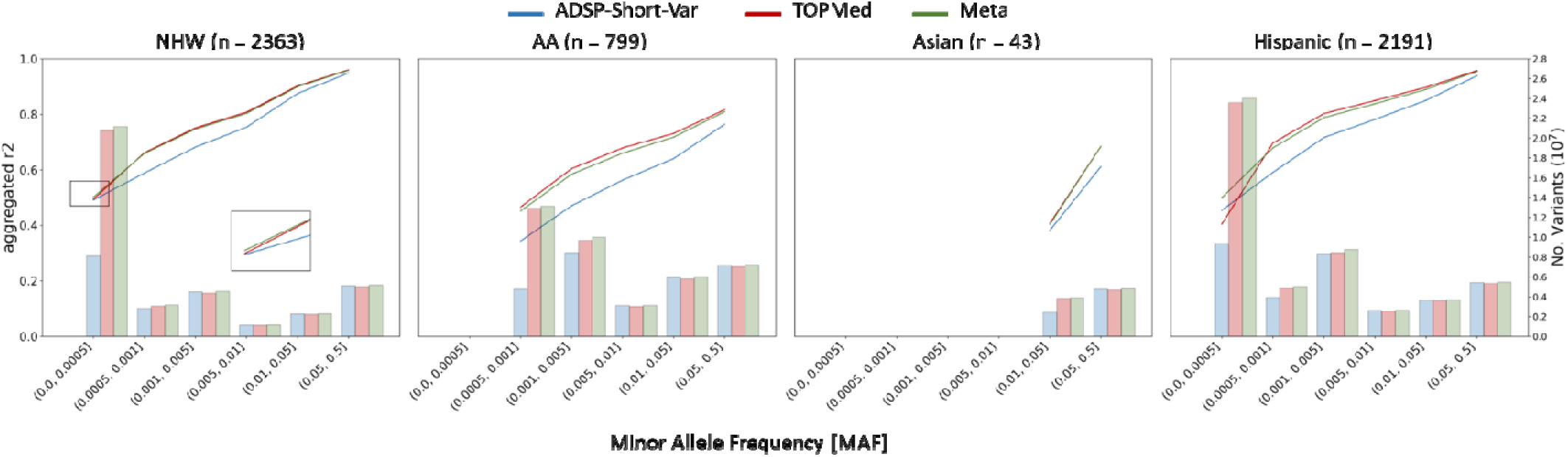
Comparison of aggregated R^2^, the squared Pearson correlation between genotypes obtained from imputations and WGS, for four ethnicities. The bars indicate the total number of variants analyzed for each ethnicity.

Using the merge feature of Meta-Minimac2^24^, a sophisticated algorithm designed to merge imputations from multiple reference panels into a unified imputation, we obtained a meta-imputation by merging imputations from the ADSP-Short-Var and TOPMed-r2 panels. The meta-imputation improved the imputation quality for variants with MAF < 0.005, elevating aggregated R^2^ from 0.4542 and 0.4045 (ADSP-Short-Var and TOPMed-r2; difference 0.0497) to 0.4995 (meta-imputation) for Hispanic cohorts and from 0.4911 and 0.4935 (difference 0.0024) to 0.5014 for NHW cohorts.

Our analyses, however, revealed a nuanced performance landscape. While Meta-Minimac2 generally improved imputation quality when initial accuracies were closely matched, such as in the MAF < 0.0005 bin for NHW cohorts, divergent outcomes were observed where substantial disparities existed between the initial imputations. Specifically, the differences in the average aggregated R² for MAFs > 0.005 were 0.0471±0.0269 for NHW, 0.1031±0.0324 for AA, 0.0492±0.0340 for Asian, and 0.0652±0.0340 for Hispanic cohorts, indicating decreased performance in these scenarios. These findings underscore the potential of tailored reference panels in improving the imputation of rare SNVs and indels and demonstrate that combining the strengths of different panels through Meta-Minimac2 can optimize imputations.

### Assessment of Imputation Accuracy of SVs Compared to WGS

Since aggRsquare is not well-suited for accurately assessing SVs, we calculated the precision and recall of SVs obtained from imputation compared to those identified by Manta on WGS for each sample, using the same dataset utilized for SNV and indel imputation accuracy. We filtered SVs identified from WGS data with the label “PASS”, and for imputed SVs, we selected the high-quality SVs listed in the previous study. Given that imputation quality (R²) is indicative of imputation accuracy, we applied different R thresholds (i.e., 0.2, 0.5, and 0.8) to filter the imputed SVs and compared them to SVs identified from WGS data. On average, there were 8,523.65±803.92 SVs per sample on WGS, whereas high-quality imputed SVs on genotype array data is on average 11211.25±303.14, 10461.67±298.75, 9696.10±290.25 and 5,418.22±242.01 for R^2^ filter set at 0, 0.2, 0.5 and 0.8 (**Figure S5**).

Both false positive (FP) and true positive (TP) SVs decreased as the R threshold increased (**Table S7**). For FPs, the numbers of deletions, duplications, and insertions dropped from 2,693.80±171.58, 282.69±23.71, and 2,199.62±197.65 with R^2^ > 0.5 to 899.42±87.26, 84.00±12.01, and 1,368.21±156.96 with R^2^ > 0.8, respectively. For TPs, the number of deletions, duplications, and insertions dropped from 2,829.95±150.55, 171.90±16.47, and 1,510.85±157.17 with R^2^ > 0.5 to 1,864.14±91.49, 90.50±10.31, and 1,104.55±118.25 with R^2^ > 0.8, respectively (**Figure 3A**). We noted that FPs dropped faster than TPs as R^2^ increased, thereby improving precision (TP / (TP + FP)). Deletions overall showed the higher precision among SV types. Specifically, average precisions were 0.6747±0.0267 for deletions, 0.5194±0.0441 for duplications, and 0.4473±0.0455 for insertions.

**Figure 3.**
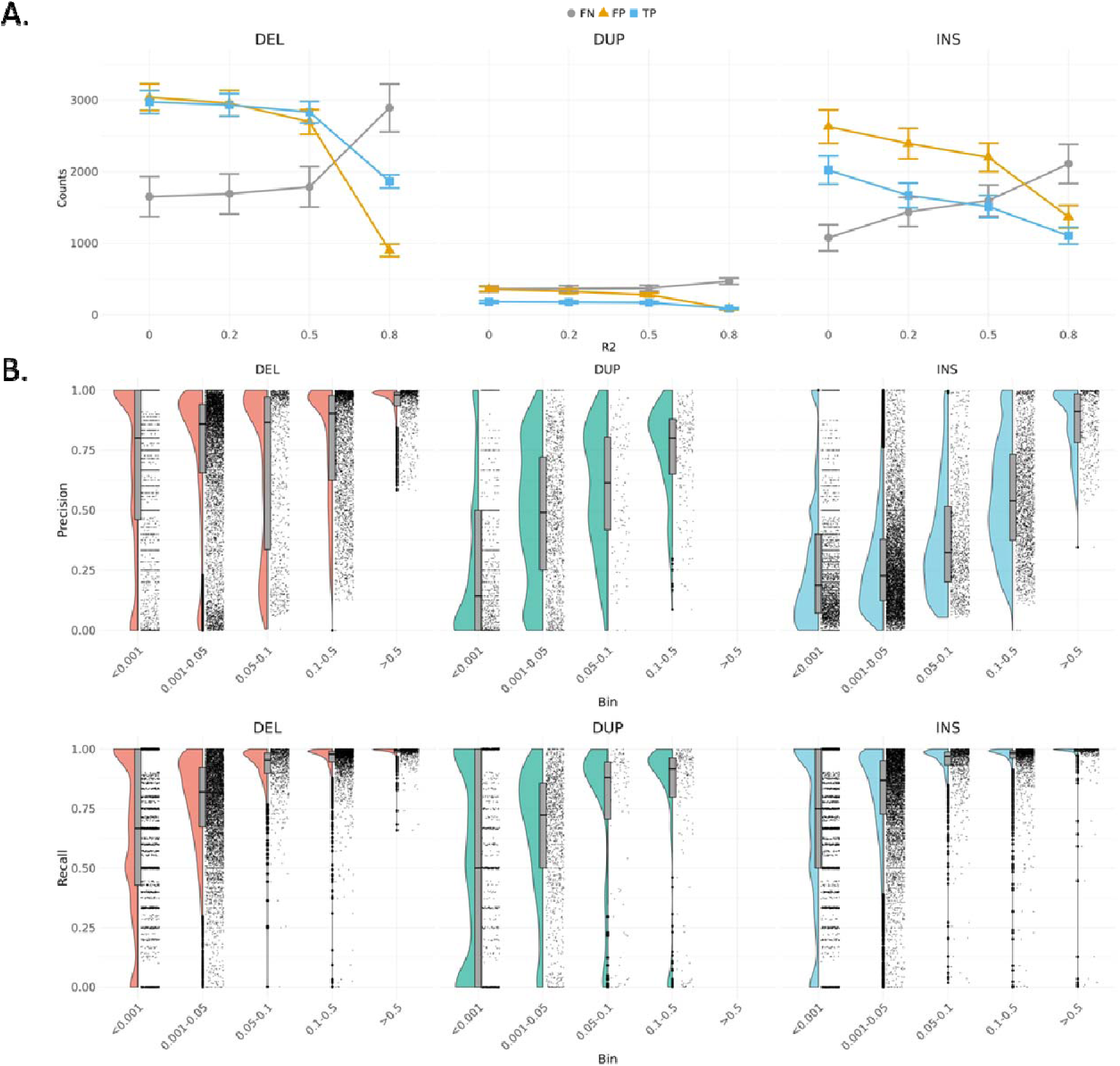
**A.** The changes of true positive, false positive and false negative among different R^2^ threshold. **B.** The precision and recall of deletions, duplications, and insertions across different allele frequency.

Conversely, the numbers of false negative (FN) increased substantially as R^2^ increases. FNs rose from 1,786.66±282.19, 372.36±40.11, and 1,596.07±217.25 with R^2^ > 0.5 to 2,890.00±332.61, 469.52±44.31, and 2,107.35±273.75 with R^2^ > 0.8 for deletions, duplications, and insertions, respectively (**Figure 3A**). Deletions stood out again with higher recall (TP / (TP + FN)) compared to other SV types, with average recalls of 0.3936±0.0204 for deletions, 0.1619±0.0149 for duplications, and 0.3446±0.0166 for insertions (**Figure S6**). The higher precision and recall of deletions reflect the better calling quality of deletions on short-read WGS compared to other types of SVs. To exhibit high accurate SVs for downstream association analysis and functional validation, we set the threshold at R^2^ > 0.8 to filter imputed SVs to obtain more promising outcomes in the analysis.

Regarding AF, we observed outstanding performance in imputing SVs with higher AF (**Figure 3B**). The average precisions were 0.9430±0.0833 for deletions and 0.8677±0.1318 for insertions with MAF > 0.5. Notice that we did not find any duplications with MAF > 0.5. Deletions maintained higher average precision in all AF bins, even at MAF < 0.001 (0.6742±0.3618). In contrast, precisions of insertions decreased rapidly from MAF > 0.5 (0.8677±0.1318) to 0.1 < MAF < 0.5 (0.5563±0.2330). Similarly, precisions of duplications dropped gradually as the AF decreased. Both deletions and insertions remained with high average recall at 0.05 < MAF < 0.1, with average recall values of 0.9233±0.0963 for deletions, 0.9337±0.1164 for insertions, and 0.7275±0.3301 for duplications. Further investigation revealed that the performance of imputing SVs is poorly correlated with SV lengths (**Figure S7**).

### Association Analyses on Different Imputations

In the ADGC genotype dataset (Ncase=16,779 and Ncontrol=21,492), we conducted single variant association tests on three distinct imputations, derived from the ADSP-Short-Var, TOPMed-r2, and meta-imputation generated by Meta-Minimac2, to evaluate the impact of various reference panels. Our analysis on pooled samples across ethnicities revealed eight genome-wide significant loci, including 495 genome-wide significant variants across 36 genes, that were concordant in all three association tests (**Table S8**). Regarding discordant signals, we found 32, 16, and 71 genome-wide significant variants uniquely identified in the ADSP-Short-Var, TOPMed-r2, and meta-imputation tests, respectively (**Figure 4, Figure S9A**). Notice that no novel significant locus was identified by those discordant variants. Furthermore, 18 suggestive significant variants in the tests on imputations from the ADSP-Short-Var (average p-value 9.76E-08±4.37E-08) and TOPMed-r2 (average p-value 6.85E-08±2.03E-08) panels showed increased significance in the test on meta-imputation (average p-value 3.86E-08±9.90E-09).

**Figure 4.**
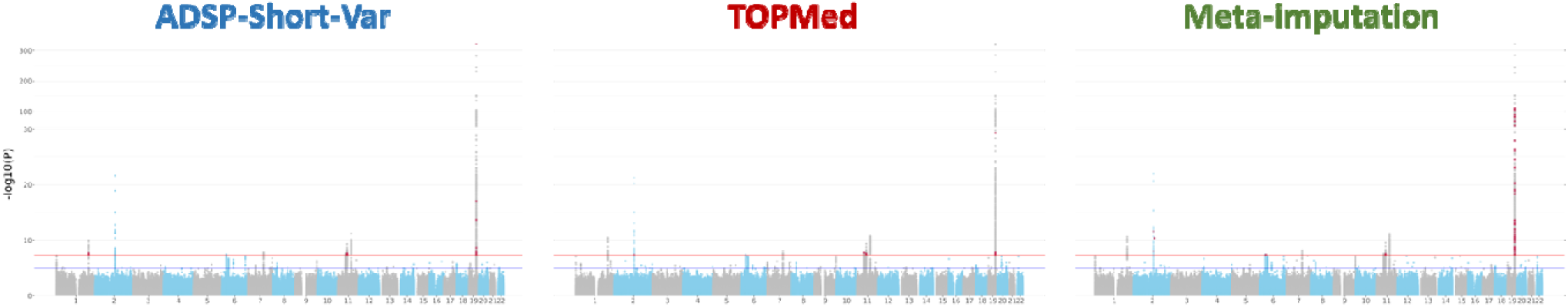
Single variant association tests performed on different imputations against the ADSP-Short-Var and TOPMed-r2 panels and meta-imputation. Red dots represent the genome-wide significant signals uniquely present in each imputation.

Ethnicity-specific analysis in NHW cohorts (Ncase=13,182 and Ncontrol=15,216) revealed eight genome-wide significant loci, including 765 genome-wide significant variants across 44 genes, in all three association tests (**Table S8**). Six of these loci, located in chromosomes 1, 2, 11, and 19, were also identified in the pooled-sample analysis. Two loci on chromosomes 1 and 3 emerged, suggesting that the approach for pooled samples might obscure specific genetic signals due to differences in population genetic structures (**Table S9**). For the panel-specific genome-wide significant variants, there were unique 36, 29, and 79 genome-wide significant variants from the ADSP-Short-Var, TOPMed-r2 and meta-imputation tests, respectively (**Figures S8A,S9B**). Most of the panel-specific suggestive significant variants were at the borderline of genome-wide significance.

In AA cohorts (Ncase=1,795 and Ncontrol=3,784), three consensus loci, including 45 variants in nine genes at chromosomes 19, 21, and 22, were identified in all three association tests. An additional genome-wide significant locus with 15 significant variants in AC019063.4 on chromosome 7 was detected in the TOPMed-r2 imputation, but the locus vanished after meta-imputation. We also found that ADSP-Short-Var panel had better imputation quality (R = 0.882±0.0288) than TOPMed (R = 0.865±0.0415) for the 15 genome-wide significant variants on chromosome 7. This finding showed that the ADSP-Short-Var panel could help refine the imputation results through meta-imputation. The number of variants unique to each test was 7 for ADSP-Short-Var, 21 for TOPMed-r2, and 14 for meta-imputation (**Figures S8B** and **S9C**).

No genome-wide significant loci were observed in the Asian (Ncase=1,576 and Ncontrol=1,951) and Hispanic (Ncase=226 and Ncontrol=541) cohorts, except for APOE-ε4 SNV rs429358, which was observed genome-wide significantly associated with AD in the TOPMed-r2 test (**Figures S8C-D** and **S9D-E**). Overall, the results of single variant association tests indicated that the ADSP-Short-Var and TOPMed-r2 imputations were largely similar. Enhancing suggestive significant signals demonstrated the potential for optimizing imputation results through meta-imputation.

### Investigation of the Impact of SV Integration in Reference Panel

To evaluate the impact of integrating SVs into the ADSP-Short-Var panel for the ADSP-All-Var panel, we assessed the imputation accuracy of these two panels. We utilized a dataset containing samples with both genotype and WGS data available, performing imputations against the ADSP-Short-Var and ADSP-All-Var panels separately. The imputation accuracy was assessed by calculating aggregated R^2^ values between SNVs from imputations and WGS, revealing nearly identical imputation accuracies (**Figure S10**).

Upon comparing the results of single variant association tests on pooled samples (Ncase=16,779 and Ncontrol=21,492) imputations from the ADSP-Short-Var and ADSP-All-Var panels, we identified eight consensus genome-wide significant loci across 36 genes (**Table S8**) in both tests (**Figure 5)**. In details, there were 75 and 139 genome-wide significant variants uniquely from ADSP-Short-Var and ADSP-All-Var, respectively. For the ADSP-All-Var specific variants, 93.53% (130 out of 139) were located in the eight consensus genome-wide significant loci. Two additional genome-wide significant loci were discovered in chromosomes 3 and 22 from the imputation against the ADSP-All-Var panel.

**Figure 5.**
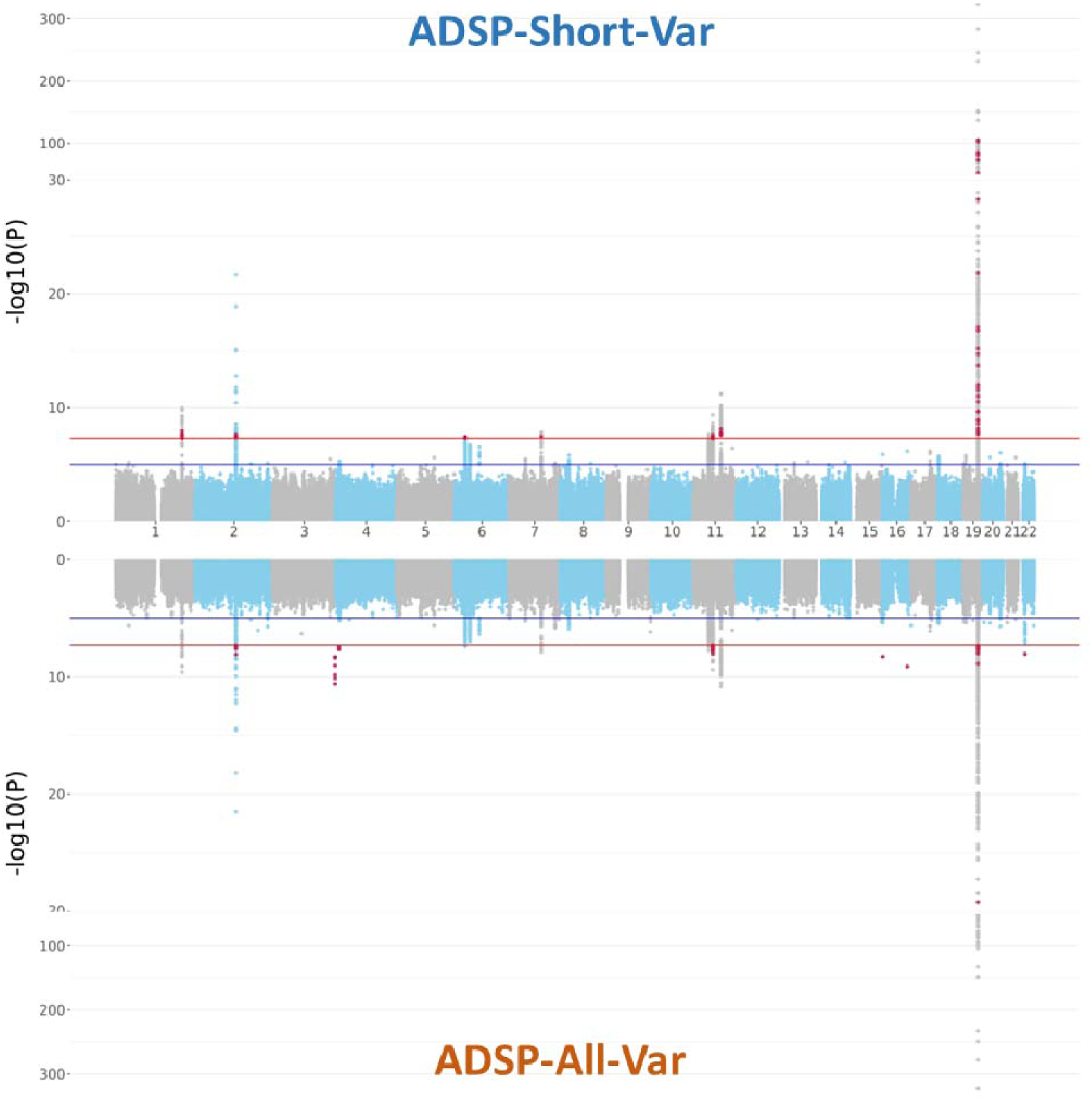
Single variant association test of imputations performed by ADSP-Short-Var and ADSP-All-Var reference panels. Red dots represent the genome-wide significant signals uniquely present in each imputation.

Compared to the ADSP-Short-Var panel, we observed that the ADSP-All-Var panel altered 8.30% of haplotypes defined by the six genome-wide significant variants on chromosome 3 and 3.06% of haplotypes defined by one genome-wide significant variant along with 48 suggestive significant variants on chromosome 22. These changes may result in differing outcomes when the same covariates are adjusted in association tests between imputations from the two panels, despite the AFs being quite similar (**Table S10**). Additionally, all the signals on chromosome 3 were within the same LD block, while all the signals on chromosome 22 were within the same LD block (**Figure S11**). Including SVs into SNV and indel panel did not dramatically alter LD structure but might have caused part of the haplotypes to change while inferring haplotypes from genotype data.

## Discussion

In this study, we constructed a reference panel of 16,564 whole-genome sequenced genomes from ADSP R3 with diverse populations including NHW, AA, Asian, and Hispanic to provide high-quality reference panels for ADRD research. To assess the performance of our reference panels, we performed imputation on ADGC datasets and compared to it from the public reference panel, TOPMed-r2. Our panel captured several rare but potential causal indels that were missed by TOPMed-r2. Furthermore, imputation from our panel provided high-quality SVs that were absent in TOPMed-r2.

We identified 3 suggestive AD associated SVs that located in two genes, *EXOC3L2* and *DMPK*. *EXOC3L2* is a component of the exocyst complex, involved in the regulation of the readily releasable pool of synaptic vesicles via the binding of NSF and SNARE proteins^26^. A variant rs597668 near *EXOC3L2* is identified as a risk factor for AD in European population^27^, but played a protective role in AD in East Asian population^28^ in previously studies. The suggestive AD associated SV in *EXOC3L2* that we identified was also recognized as a risk factor in the NHW population and as a protective factor in the Asian population. DMPK is a serine/threonine kinase that could prevent ROS-induced cell death^29^, and its gene mutations cause myotonic dystrophy type 1 (DM1)^30^. A study indicated that the expression of *DMPK* in the brain follows an age-related pattern^31^, but its role in aging or in AD is still unknown.

There were 10 out of 55 rare indels absent in TOPMed-r2 imputation, which are located in genes, *C12orf81*, *TOMM20L*, *FAM174B*, *NTN3*, *RGL3*, *PNKP*, *PPDPF*, and *PCDHB13*. *NTN3* (netrin-3) is a member of the netrin family, a kind of extracellular protein that directs cell and axon migration during embryogenesis^32^ and is highly expressed in sensory ganglia^33^. This protein family includes the other famous protein netrin-1 which is highly correlated with Aβ levels in the brain tissue of AD patients^34–36^. *PNKP* (Polynucleotide Kinase-Phosphatase) is involved in DNA repair processing^37^, that might associate with AD pathogenesis and its dysfunction of this gene can result in microcephaly or neurodegeneration^38^. The *PPDPF* was predicted to be involved in cell differentiation and mainly expressed in oligodendrocytes based on the data from the human protein atlas. It was downregulated in the dorsolateral prefrontal cortex of AD patients comparing to people with NCI (no cognitive impairment) or MCI (mild cognitive impairment)^39^.

Each reference panel offers unique strengths, leading us to employ Meta-Minimac2 for meta-imputation. This tool integrates imputations from multiple reference panels, leveraging their collective strengths to enhance imputation accuracy. Our application of Meta-Minimac2 improved imputation results for ultra-rare variants in the NHW and Hispanic ethnic groups. However, in AA and Asian groups, the accuracy of meta-imputation was not improved. This suggests that meta-imputation does not universally enhance accuracy, particularly when substantial discrepancies exist between initial imputation results. Despite these challenges, meta-imputation still enabled the identification of AD-specific genotypes absent in TOPMed-r2. From single variant association tests, we found that most genome-wide significant signals were consistent across imputations from the ADSP-Short-Var and TOPMed-r2 panels, and through meta-imputation. However, meta-imputation introduced some noise signals, which lacks LD support.

In this study, we expanded our methodologies by constructing a reference panel integrated with SVs. Previous research has applied similar approaches to impute SVs for general populations and cardiometabolic traits^40,41^; however, these studies did not specifically address AD. Efforts have been made to use high-quality SV datasets to improve SV imputation on genotype data. Our inclusion of high-quality SVs into the imputation panel resulted in commendable imputation quality. Nevertheless, the existing pipeline fails to address potential conflicts among SNVs, indels, and SVs. For example, SNVs should not be present within homozygous deletions. This oversight indicates a clear necessity for novel phasing or imputation methods specifically designed for SVs. Ultimately, our tailored reference panel promises to significantly advance genetics research in Alzheimer’s Disease and Related Dementias (ADRD), especially concerning rare variants and SVs.

## Materials and Methods

### Whole Genome Sequence Samples From ADSP

Alzheimer’s Disease Sequencing Project (ADSP)^42^ is a collaborative project aiming at identifying new variants, genes, and therapeutic targets in AD. Data from the ADSP are available to qualified investigators via the National Institute on Aging Genetics of Alzheimer’s Disease Data Storage Site (NIAGADS) (https://dss.niagads.org/). This work focused on participants with WGS in the NIAGADS data named “R3 17K WGS Project Level VCF”, which contains 16,905 subjects (6,646 AD cases, 6,938 controls and 3,321 subjects with unknown AD status) collected across 24 cohorts and whole genome sequencing was performed by Illumina HiSeqX, HiSeq2000, HiSeq2500, and NovaSeq platforms. The ADSP dataset included 10,517 non-Hispanic white, 3,018 African American, 3,296 Hispanic white, 50 Asian and 24 unknown or other ethnicities.

From the initial pool of 16,905 individuals of the ADSP R3 17K, we first removed 341 related samples through the identity by descent (IBD) analysis (PI_HAT > 0.4). Subsequently, we conducted a rigorous variant quality control process, starting with the assessment of Hardy-Weinberg Equilibrium (HWE) using RUTH^43^ with the top 10 principal components (PCs) that calculate by plink in control subjects and filtering variants violating the principle (SLE_P_I < −4). Variants with an allele count (AC) less than 5 and a missing genotype rate exceeding 90% were also removed. This stringent filtering resulted in a final dataset of 16,564 sequenced genomes with 51,459,037 SNVs and 3,322,380 indels for a reference panel construction (**Figure S12A**).

### Genotype Array Samples From ADGC

The Alzheimer’s Disease Genetics Consortium (ADGC) is a collection of GWAS data funded by the NIH, aiming to collaboratively use the collective resources of the AD research community to resolve Alzheimer’s disease (AD) genetics. In total, 51 available cohorts with 21,492 control and 16,779 AD cases were used in this study. There were 15,176 male and 23,095 female in this dataset. This dataset consists of 4 ethnicities that include 28,398 non-Hispanic white, 5,579 African American, 3,527 Asian and 767 Hispanic samples.

### Whole Genome Sequence Data Process

The Genome Center for Alzheimer’s Disease (GCAD) mapped short reads against the reference genome hg38 using BWA MEM^44^, called SNVs and indels using the GATK HaplotypeCaller V2.6^45^ for each sample, and then performed joint genotyping across all samples using GATK. The GCAD quality control (QC) working group performed quality checks of variants and genotypes and assigned a quality annotation^46^.

The SV callset is available on NIAGADS as well^17^ For each sample, Manta^25^ v1.6.0 and Smoove v0.2.5 (https://github.com/brentp/smoove) with default parameters were used. Calls from Manta and Smoove were merged by Svimmer^47^ to generate a union of two call sets for a sample. Unresolved non-reference ‘breakends’ (BNDs) and SVs > 10 Mb were filtered. Then, all individual sample VCF files were merged together by Svimmer as input to Graphtyper2^48^ v2.7.3 for joint genotyping. This study utilizes the SVs from the callset^17^.

### Workflow of Reference Panel Building

We first phased 51,459,037 SNVs and 3,322,380 indels derived from the 16,564 whole genome sequenced genomes by SHAPEIT4^49^, followed by converting the vcf format into m3vcf using minimac3^50^. At last, we could get the ADSP-Short-Var panel. In order to extend our research into SVs, we augmented our reference panel to include not only SNVs and indels but also SVs. Leveraging SV callset obtained from our previous study^17^ on ADSP R3 WGS data, we selected and incorporated 231,385 deletions, 119,648 insertions, 45,839 duplications and 3,362 inversions were selected and incorporated into ADSP-Short-Var panel construction. The high-quality SVs were merged with a stringent filtered ADSP 17K dataset. Then phased the dataset, which contained SNVs, indels, and SVs, by SHAPEIT4 and then turned the vcf format into m3vcf by minimac3^50^. After this process, we obtained the ADSP-All-Var panel.

### Workflow of Genotype Imputation

The imputation strategy was shown in **Figure S12B**. Total 51 ADGC cohort was first phased by SHAPEIT4-4.2.2^49^ and imputed on the TOPMed imputation server^6^ by TOPMed-r2 panel. The phased datasets also imputed to ADSP-Short-Var panel and ADSP-All-Var panel by minimac4-1.0.2^50^. Then we utilized metaminimac2-1.0.0^24^ to combine imputation results generated using TOPMed and ADSP-Short-Var panel into a consensus imputed dataset. To merge all imputed cohorts of each imputation, the imputation quality scores (R^2^) were calculated and combined using Fisher z-transformation and generated lists of excluded and retained variants from information files (.info.gz) by IMMerge^51^. We removed SNVs which R^2^ labeled as NA in information files that were generated by TOPMed imputation server in order to avoid the failed calculation of Fisher z-transformation before the merging process start. At last, 38,271 samples with known AD status were selected from the merged cohort to form the final dataset.

### Single variant association analysis

For the single variant association test, variants with R^2^ over 0.8, MAF over 0.5%, and the HWE_SLP_I value range from −4 to 4 were used in the task. We used a R package GENESIS^52^ v. 2.28.0 to perform single variant and structure variant association test with an additive genotype model adjusting for age, sex and population substructure using top 10 principal components.

### Imputation accuracy and quality measurement

Imputation accuracy was determined by comparing genotypes from imputation to genotypes from WGS. The WGS data of 36,361 individuals of the ADSP R4 36K were utilized to evaluate the imputation accuracy of imputed genotypes. The variants were called by GATK^45^ v.4.1.1 and SVs were generated by manta. We utilized 5396 samples (2363 non-Hispanic white, 2191 Hispanic, 799 African American, 43 Asian) which both had genotype array data and WGS data, independent to samples used in building panel, for evaluating the imputation accuracy. The validation samples were selected from each imputed cohort and merge together by ethnicity using bcftools^53^ for the three imputations. For each ethnicity, all three imputations were compared to WGS data through calculating aggregated r2 by aggRsquare^24^, which is calculated as the squared Pearson correlation between the imputed genotypes and the WGS genotypes. Imputation quality was determined by R^2^ score, that generated from Minimac4. The threshold of well-imputed variants was setting at the R^2^ over 0.8.

Imputed SVs with R^2^ over 0.8 were kept for validation. SVs callset were from Manta^25^. The same SVs were defined by the covered region of each structure variant in imputed SVs reciprocal overlap more than 50% with the SVs in validation call set. BEDTools^54^ was used to intersect the SVs. For each sample, the SVs discovered in both imputation and validation dataset were deemed as true positive. The SVs only discovered in imputation dataset were defined as false positive, in contrast, the SVs only discovered in validation dataset were defined as false negative. The precision of each SVs was calculated by the number of all true positive in the SV divide the sum of the number of the true positive and the number of false positive in the same SV. On the other hand, the recall of each SVs was calculated by the number of all true positive in the SV divide the sum of the number of the true positive and the number of false negative in the same SV. The allele frequency of validation dataset was used to assign SVs to specific allele frequency bin.

### PCR validation

For variant’s genotyping, primers were designed at 200bp upstream and downstream of the target position. 50ng Genomic DNA was amplified by SimpliAmp Thermal Cycler (Applied Biosystems) in a 20ul reaction volume with HotStarTaq Master Mix (Qiagen) in the presence of 2uM primers (IDT). PCR was performed at: 95°C for 15min; 30 cycles at 95°C for 20sec, 55°C for 30sec, 72°C for 2min; with a final extension of 72°C for 7min. The amplified target sequences were cleaned up with ExoSAP-IT (USB) by incubating at 37°C for 45min followed by 80°C for 15min. The target sequences after being cleaned up, were then used to perform Sanger sequencing by using the BigDye® Terminator v3.1 Cycle Sequencing kit (Part No. 4336917 Applied Biosystems) at: 96°C for 1min; 25 cycles at 96°C for 10sec, 50°C for 5sec, 60°C for 1min15sec. The products were then cleaned up by using XTerminator and SAM Solution (Applied Biosystems) with 30min of shaking at 1800rpm. The sequencing products were analyzed on a SeqStudio Genetic Analyzer (Applied Biosystems) and the sequencing traces were analyzed using Sequencher 5.4 (Gene Code).

### Statistical analysis

For comparing the rare variants and SVs of ADSP dataset to imputations of ADSP, associations of case and control were calculated by Fischer’s exact test. Pearson correlation was used to estimate the correlation of AFs between ADSP-Short-Var imputation and AFs discovered from ADSP R3 WGS data. A P value that less than 0.05 was determined as nominal significant. All statistical analyses were performed in R.

## Acknowledgements

See supplementary text. PLC, HW, IH, and TC report grant support from RF1-AG074328. WPL reports grant support from RF1-AG074328, P30-AG072979, U54-AG052427, and U24-AG041689.

## Author contributions

PLC, HW, and IH performed statistical analyses. PLC and HW performed phenotype acquisition and/or harmonization. PLC, HW, and WPL performed Genotype acquisition and/or QC. BAD, PLC, and GDS performed experimental validation. PLC, HW, JJF, IH, TC, GT, BWK, WSB, BV, GDS, and WPL interpretated results. PLC and WPL wrote the first draft of the manuscript. All authors read, critically revised, and approved the manuscript.

## Data availability

https://github.com/whtop/SV-ADSP-Pipeline https://dss.niagads.org/

## Code availability

ADSP-Short-Var and ADSP-All-Var panel building codes are publicly accessible at https://github.com/plCas/SNP-SV-imputation-panel-building-pipeline

## Reference

1 International HapMap, C., et al. Integrating common and rare genetic variation in diverse human populations. Nature 467, 52–58 (2010). 10.1038/nature09298

2 Genomes Project, C., et al. A map of human genome variation from population-scale sequencing. Nature 467, 1061–1073 (2010). 10.1038/nature09534

3 Huang, J. et al. Improved imputation of low-frequency and rare variants using the UK10K haplotype reference panel. Nat Commun 6, 8111 (2015). 10.1038/ncomms9111

4 McCarthy, S. et al. A reference panel of 64,976 haplotypes for genotype imputation. Nat Genet 48, 1279–1283 (2016). 10.1038/ng.3643

5 Bick, A. G. et al. Inherited causes of clonal haematopoiesis in 97,691 whole genomes. Nature 586, 763–768 (2020). 10.1038/s41586-020-2819-2

6 Taliun, D. et al. Sequencing of 53,831 diverse genomes from the NHLBI TOPMed Program. Nature 590, 290–299 (2021). 10.1038/s41586-021-03205-y

7 Choi, J. et al. A whole-genome reference panel of 14,393 individuals for East Asian populations accelerates discovery of rare functional variants. Sci Adv 9, eadg6319 (2023). 10.1126/sciadv.adg6319

8 O’Connell, J. et al. A population-specific reference panel for improved genotype imputation in African Americans. Commun Biol 4, 1269 (2021). 10.1038/s42003-021-02777-9

9 Girirajan, S. et al. Relative burden of large CNVs on a range of neurodevelopmental phenotypes. PLoS Genet 7, e1002334 (2011). 10.1371/journal.pgen.1002334

10 de Cid, R. et al. Deletion of the late cornified envelope LCE3B and LCE3C genes as a susceptibility factor for psoriasis. Nat Genet 41, 211–215 (2009). 10.1038/ng.313

11 Allen, M. et al. Association of MAPT haplotypes with Alzheimer’s disease risk and MAPT brain gene expression levels. Alzheimers Res Ther 6, 39 (2014). 10.1186/alzrt268

12 Brouwers, N. et al. Alzheimer risk associated with a copy number variation in the complement receptor 1 increasing C3b/C4b binding sites. Mol Psychiatry 17, 223–233 (2012). 10.1038/mp.2011.24

13 Li, Y. et al. Integrated copy number and gene expression analysis detects a CREB1 association with Alzheimer’s disease. Transl Psychiatry 2, e192 (2012). 10.1038/tp.2012.119

14 Noyvert, B. et al. Imputation of structural variants using a multi-ancestry long-read sequencing panel enables identification of disease associations. medRxiv, 2023.2012.2020.23300308 (2023). 10.1101/2023.12.20.23300308

15 Langlois, A. W. R. et al. Genotyping, characterization, and imputation of known and novel CYP2A6 structural variants using SNP array data. J Hum Genet 68, 533–541 (2023). 10.1038/s10038-023-01148-y

16 McLaren, W. et al. The Ensembl Variant Effect Predictor. Genome Biol 17, 122 (2016). 10.1186/s13059-016-0974-4

17 Wang, H. et al. Structural Variation Detection and Association Analysis of Whole-Genome-Sequence Data from 16,905 Alzheimer’s Diseases Sequencing Project Subjects. medRxiv (2023). 10.1101/2023.09.13.23295505

18 Schwartzentruber, J. et al. Genome-wide meta-analysis, fine-mapping and integrative prioritization implicate new Alzheimer’s disease risk genes. Nat Genet 53, 392–402 (2021). 10.1038/s41588-020-00776-w

19 Paul, V. et al. Scratch2 modulates neurogenesis and cell migration through antagonism of bHLH proteins in the developing neocortex. Cereb Cortex 24, 754–772 (2014). 10.1093/cercor/bhs356

20 Itoh, Y. et al. Scratch regulates neuronal migration onset via an epithelial-mesenchymal transition-like mechanism. Nat Neurosci 16, 416–425 (2013). 10.1038/nn.3336

21 Rodriguez-Aznar, E. & Nieto, M. A. Repression of Puma by scratch2 is required for neuronal survival during embryonic development. Cell Death Differ 18, 1196–1207 (2011). 10.1038/cdd.2010.190

22 Schenning, K. J. et al. Gene-Specific DNA Methylation Linked to Postoperative Cognitive Dysfunction in Apolipoprotein E3 and E4 Mice. J Alzheimers Dis 83, 1251–1268 (2021). 10.3233/JAD-210499

23 Lee, W. P. et al. Association of Common and Rare Variants with Alzheimer’s Disease in over 13,000 Diverse Individuals with Whole-Genome Sequencing from the Alzheimer’s Disease Sequencing Project. medRxiv (2023). 10.1101/2023.09.01.23294953

24 Yu, K. et al. Meta-imputation: An efficient method to combine genotype data after imputation with multiple reference panels. Am J Hum Genet 109, 1007–1015 (2022). 10.1016/j.ajhg.2022.04.002

25 Chen, X., et al. Manta: rapid detection of structural variants and indels for germline and cancer sequencing applications. Bioinformatics 32, 1220–1222 (2016). 10.1093/bioinformatics/btv710

26 Robbins, M., Clayton, E. & Kaminski Schierle, G. S. Synaptic tau: A pathological or physiological phenomenon? Acta Neuropathol Commun 9, 149 (2021). 10.1186/s40478-021-01246-y

27 Seshadri, S. et al. Genome-wide analysis of genetic loci associated with Alzheimer disease. JAMA 303, 1832–1840 (2010). 10.1001/jama.2010.574

28 Wu, Q. J. et al. EXOC3L2 rs597668 variant contributes to Alzheimer’s disease susceptibility in Asian population. Oncotarget 8, 20086–20091 (2017). 10.18632/oncotarget.15380

29 Pantic, B. et al. Myotonic dystrophy protein kinase (DMPK) prevents ROS-induced cell death by assembling a hexokinase II-Src complex on the mitochondrial surface. Cell Death Dis 4, e858 (2013). 10.1038/cddis.2013.385

30 Kaliman, P. & Llagostera, E. Myotonic dystrophy protein kinase (DMPK) and its role in the pathogenesis of myotonic dystrophy 1. Cell Signal 20, 1935–1941 (2008). 10.1016/j.cellsig.2008.05.005

31 Langbehn, K. E. et al. DMPK mRNA Expression in Human Brain Tissue Throughout the Lifespan. Neurol Genet 7, e537 (2021). 10.1212/NXG.0000000000000537

32 Rajasekharan, S. & Kennedy, T. E. The netrin protein family. Genome Biol 10, 239 (2009). 10.1186/gb-2009-10-9-239

33 Wang, H., Copeland, N. G., Gilbert, D. J., Jenkins, N. A. & Tessier-Lavigne, M. Netrin-3, a mouse homolog of human NTN2L, is highly expressed in sensory ganglia and shows differential binding to netrin receptors. J Neurosci 19, 4938–4947 (1999). 10.1523/JNEUROSCI.19-12-04938.1999

34 Meng, Y., Sun, S., Cao, S. & Shi, B. Netrin-1: A Serum Marker Predicting Cognitive Impairment after Spinal Cord Injury. Dis Markers 2022, 1033197 (2022). 10.1155/2022/1033197

35 Ju, T. et al. Decreased Netrin-1 in Mild Cognitive Impairment and Alzheimer’s Disease Patients. Front Aging Neurosci 13, 762649 (2021). 10.3389/fnagi.2021.762649

36 Bai, B. et al. Deep Multilayer Brain Proteomics Identifies Molecular Networks in Alzheimer’s Disease Progression. Neuron 105, 975–991 e977 (2020). 10.1016/j.neuron.2019.12.015

37 Weinfeld, M., Mani, R. S., Abdou, I., Aceytuno, R. D. & Glover, J. N. Tidying up loose ends: the role of polynucleotide kinase/phosphatase in DNA strand break repair. Trends Biochem Sci 36, 262–271 (2011). 10.1016/j.tibs.2011.01.006

38 Dumitrache, L. C. & McKinnon, P. J. Polynucleotide kinase-phosphatase (PNKP) mutations and neurologic disease. Mech Ageing Dev 161, 121–129 (2017). 10.1016/j.mad.2016.04.009

39 McCorkindale, A. N., Patrick, E., Duce, J. A., Guennewig, B. & Sutherland, G. T. The Key Factors Predicting Dementia in Individuals With Alzheimer’s Disease-Type Pathology.Front Aging Neurosci 14, 831967 (2022). 10.3389/fnagi.2022.831967

40 Hehir-Kwa, J. Y. et al. A high-quality human reference panel reveals the complexity and distribution of genomic structural variants. Nat Commun 7, 12989 (2016). 10.1038/ncomms12989

41 Chen, L. et al. Association of structural variation with cardiometabolic traits in Finns. Am J Hum Genet 108, 583–596 (2021). 10.1016/j.ajhg.2021.03.008

42 Beecham, G. W. et al. The Alzheimer’s Disease Sequencing Project: Study design and sample selection. Neurol Genet 3, e194 (2017). 10.1212/NXG.0000000000000194

43 Kwong, A. M. et al. Robust, flexible, and scalable tests for Hardy-Weinberg equilibrium across diverse ancestries. Genetics 218 (2021). 10.1093/genetics/iyab044

44 Li, H. & Durbin, R. Fast and accurate short read alignment with Burrows-Wheeler transform. Bioinformatics 25, 1754–1760 (2009). 10.1093/bioinformatics/btp324

45 McKenna, A. et al. The Genome Analysis Toolkit: a MapReduce framework for analyzing next-generation DNA sequencing data. Genome Res 20, 1297–1303 (2010). 10.1101/gr.107524.110

46 Naj, A. C. et al. Quality control and integration of genotypes from two calling pipelines for whole genome sequence data in the Alzheimer’s disease sequencing project.Genomics 111, 808–818 (2019). 10.1016/j.ygeno.2018.05.004

47 GitHub-DecodeGenetics/svimmer. Structural Variant Merging Tool. (2021).

48 Eggertsson, H. P. et al. GraphTyper2 enables population-scale genotyping of structural variation using pangenome graphs. Nat Commun 10, 5402 (2019). 10.1038/s41467-019-13341-9

49 Delaneau, O., Zagury, J. F., Robinson, M. R., Marchini, J. L. & Dermitzakis, E. T. Accurate, scalable and integrative haplotype estimation. Nat Commun 10, 5436 (2019). 10.1038/s41467-019-13225-y

50 Das, S. et al. Next-generation genotype imputation service and methods. Nat Genet 48, 1284–1287 (2016). 10.1038/ng.3656

51 Zhu, W. et al. IMMerge: merging imputation data at scale. Bioinformatics 39 (2023). 10.1093/bioinformatics/btac750

52 Gogarten, S. M. et al. Genetic association testing using the GENESIS R/Bioconductor package. Bioinformatics 35, 5346–5348 (2019). 10.1093/bioinformatics/btz567

53 Danecek, P., et al. Twelve years of SAMtools and BCFtools. Gigascience 10 (2021). 10.1093/gigascience/giab008

54 Quinlan, A. R. & Hall, I. M. BEDTools: a flexible suite of utilities for comparing genomic features. Bioinformatics 26, 841–842 (2010). 10.1093/bioinformatics/btq033

